# Mapping Digital and Assistive Technologies for Dementia and Related Neurodegenerative Conditions: A Scoping Review

**DOI:** 10.64898/2026.06.30.26356906

**Authors:** Selin Birgen, Macy Fan, Sikander Chimbaiwala, Julian Jeyasingh-Jacob, Christopher Fox, Helen Dawes, Shlomi Haar

**Author notes:** Correspondence to: Shlomi Haar. SB and MF equally contributed as co-first authors. HD and SH equally contributed as co-senior authors.

## Abstract

Neurodegenerative conditions increasingly adopt digital and assistive technologies in community and home-based settings to support independence, symptom management and quality of life. This registered scoping review (https://doi.org/10.17605/OSF.IO/QJZC9) followed Joanna Briggs Institute methodology and PRISMA-ScR reporting guidelines to map the evidence base and assess technology adoption, methodological maturity and translational readiness. We identified 23,453 records published between 2001 and 2025, of which 325 studies met the inclusion criteria. Parkinson’s disease represented 49.5% of studies, followed by Alzheimer’s and other dementias (33.6%), whereas Huntington’s disease and motor neuron disease were rarely represented. The evidence base expanded rapidly over the last decade but was dominated by small, short-duration, prototype-based evaluations. Mobility and Health Monitoring comprised 73.5% of studies, while Communication and Activities of Daily Living support were comparatively underrepresented. Greater convergence around validated platforms and evaluation pathways may be required to improve translation and real-world impact.

## Introduction

Neurodegenerative conditions include dementias (Alzheimer’s disease (AD), Vascular dementia, Lewy Body dementia), and other disorders such as Parkinson’s disease (PD), motor neuron disease (MND), and Huntington’s disease (HD). These share a clinical trajectory of progressive cognitive and motor decline, communication difficulty, and reduced capacity for independent daily living^1,2^. Dementia alone affects approximately 57 million people worldwide and is projected to reach 139 million by 2050^3,4^. With disease-modifying therapies remaining limited for most of these conditions, supportive care aimed at preserving function, autonomy, and caregiver capacity has become a central priority in clinical practice and health policy^5,6^. The COVID-19 pandemic accelerated an existing shift of care from institutional to home and community settings^7^. Around two-thirds of people living with dementia reside in community settings rather than in care homes^8^. Yet, sustaining safe home-based care presents challenges, including medication errors^9^, social isolation^10,11^, and delayed recognition of health deterioration^12^, ultimately placing additional strains on individuals, families, and caregivers^13^.

Digital and assistive technologies are widely positioned to support clinical assessment and monitoring, and to promote independence, safety, and caregiver capacity in home-based care^14^. Over the past two decades, a substantial body of literature has developed around these technologies for people with neurodegenerative conditions. This work spans over five broad domains relevant to home care: mobility, health monitoring, activities of daily living, communication, and caregiver support. Within the dementia literature alone, recent reviews have reported promising effects across cognitive, health, and social outcomes when technologies are tailored to users’ needs^14^. Yet, evidence of sustained adoption in routine care has been less consistent.

Despite this scale of activity, translation into clinical practice has remained limited^15,16^. Many technologies demonstrate feasibility or short-term acceptability in early studies yet fail to progress to sustained adoption in real-world settings. This pattern is characterised in digital health implementation as ’pilotitis’^17,18^, and aligns with the Non-adoption, Abandonment, Scale-up, Spread, and Sustainability (NASSS) framework’s account of why digital health innovations stall during scale-up^19^. Contributing factors across studies include short evaluation windows, reliance on research-stage hardware, and limited end-user involvement. Previous reviews have described effectiveness and implementation barriers, but they have not quantitatively characterised how the primary literature is distributed across conditions, domains and methodological characteristics, considering pathway translation at the field level^14,20,21^.

This scoping review, commissioned by the Zero Burden Sustainable Technologies to support independent living with dementia (ZeDTech) Network, maps the evidence base for digital and assistive technologies in dementia and related neurodegenerative conditions across the five domains central to home-based care. We characterise what technologies have been studied, in which populations, with what study designs and stages, and how usability, acceptability, and adherence have been evaluated. To examine the field’s methodological maturity, we characterise the distribution of and type of evaluation effort across included studies using participant-days (the product of sample size and observation duration) and report how studies cluster against the distributions of hardware/software origin and evaluation instrument standardisation. The aim is to identify where current technologies cluster, where evidence remains limited, and whether the field has reached the methodological scale needed to support clinically translatable evidence.

## Results

### 2.1 Search Outcomes

Our systematic literature search across eight databases yielded 23,453 records. After removing 6,118 duplicates, 17,335 unique records were included for title and abstract screening, and 16,374 were excluded at this stage for not meeting our predefined eligibility criteria, such as no technology components, wrong study designs, and wrong population. Out of 961 reports sought out for full-text retrieval, 41 could not be retrieved through institutional access channels and were excluded. The remaining 920 reports were assessed for eligibility, and 595 were excluded with reasons detailed in Fig. 1, leaving 325 studies for data extraction and synthesis. Inter-rater agreement at full-text screening was high (κ = 0.97); disagreements at each stage were resolved through discussion, with arbitration by a third reviewer where consensus could not be reached.

**Figure 1.**
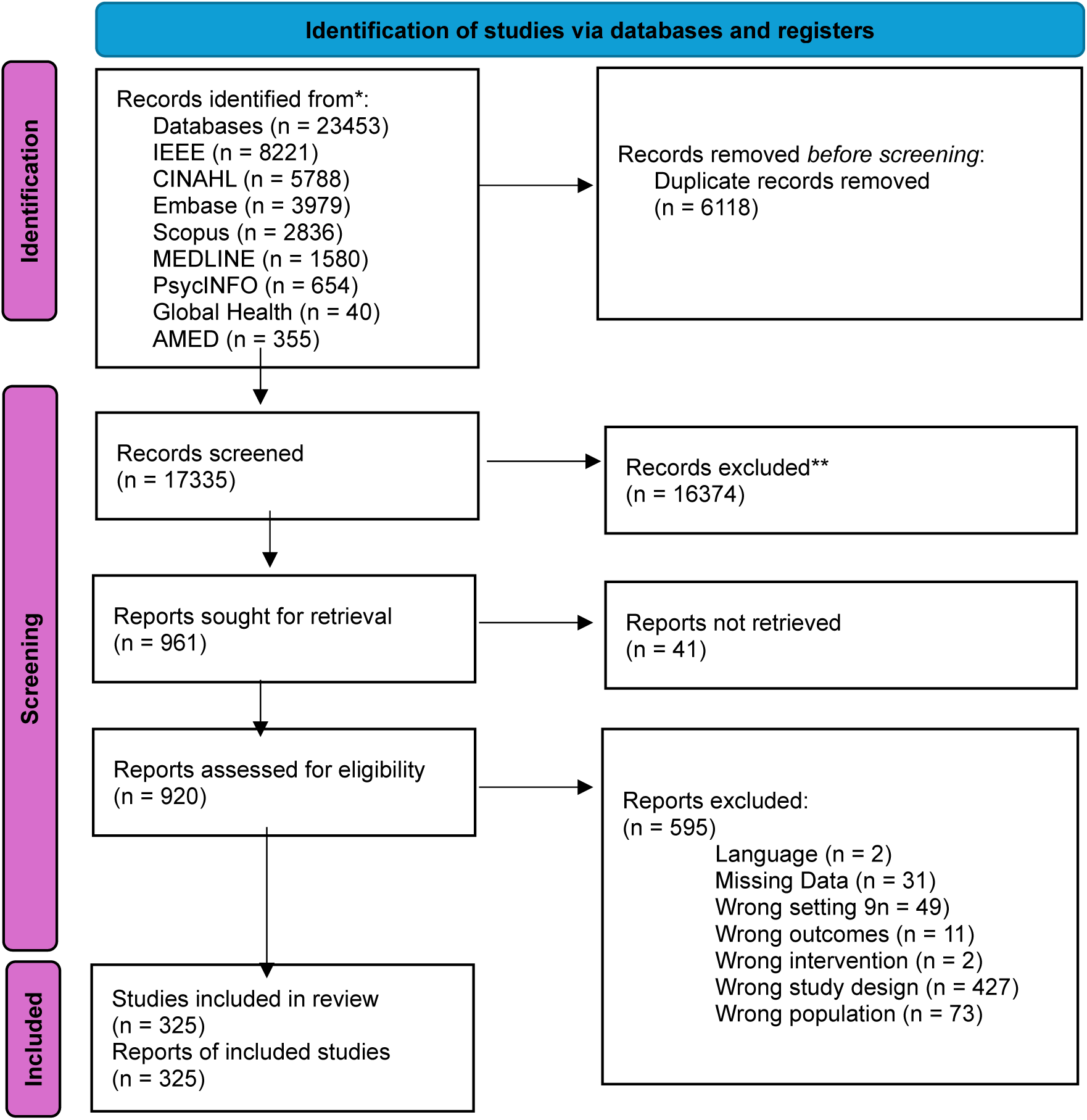
PRISMA-ScR flow diagram showing the identification, screening, eligibility assessment, and inclusion of studies. Exclusion reasons at full-text stage are shown in the corresponding box.

### 2.2 Publication Trends and Clinical Populations

Publication volume has grown rapidly in recent years, while the diagnostic distribution remains narrow (Fig. 2). Publication output across the 25-year review period accelerated substantially over time (Fig. 2A). Between 2001 and 2010, the field produced fewer than two studies per year on average, 15 studies in total across the decade. Output began to rise from 2016 onward and continued to accelerate, with 42 studies recorded in the first ten months of 2025 alone, surpassing the 2024 total (n = 36). The 195 studies published from 2020 onward exceed the entire pre-2020 record (n = 130) by 50%, despite covering less than a quarter of the review period. Mobility and Health Monitoring (blue and green bars, respectively) were the most consistently represented domains throughout the period and remain the dominant categories in every recent year. ADL Support, Caregiver Support, and Communication (red, yellow, and purple bars, respectively) appear more sporadically in the earlier years and grew more substantially in volume from 2017 onwards, while still representing a minority of the annual output even in the most recent years.

**Figure 2.**
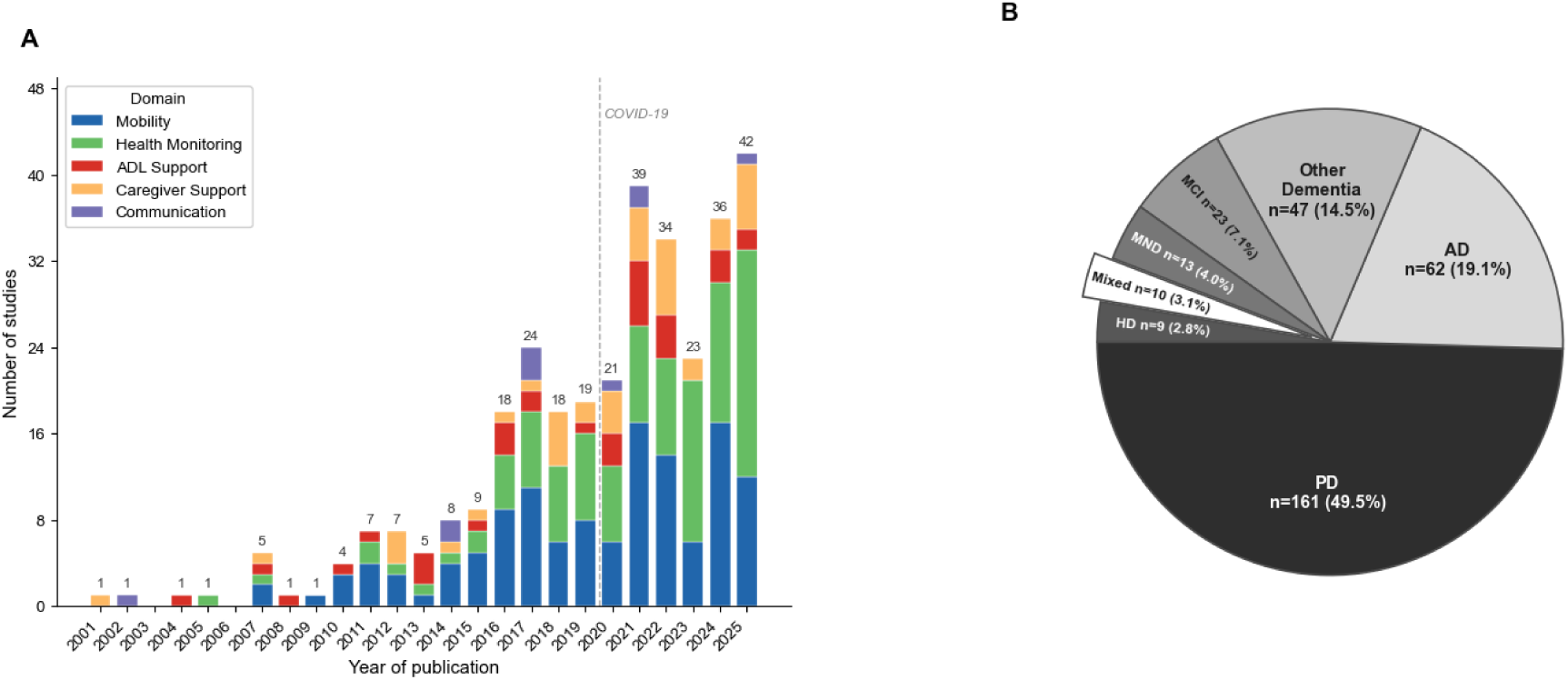
Publication trends and clinical populations of included studies. **(A)** Number of published studies per year from 2001 to 2025, stacked by primary technology domain. Dashed line indicates the onset of the COVID-19 pandemic. **(B)** Distribution of primary target diagnoses across included studies. The “Other Dementia” category comprises mixed-etiology dementia, unspecified-subtype dementia, dementia with Lewy bodies, frontotemporal dementia, Parkinson’s disease dementia, and vascular dementia. The “Mixed” category represents studies enrolling mixed neurodegenerative populations that could not be assigned to a single diagnostic category. Slices are shaded on a greyscale gradient from darkest (PD) to lightest (AD), with Mixed populations shown in white and exploded. *Abbreviations: ADL, Activities of Daily Living; PD, Parkinson’s Disease; AD, Alzheimer’s Disease; HD, Huntington’s Disease; MCI, Mild Cognitive Impairment; MND, Motor Neuron Disease*.

Alongside this growth, the distribution of primary target diagnoses was narrow (Fig. 2B). Of the 325 included studies, Parkinson’s disease (PD) was the most frequently studied condition (n = 161, 49.5%), followed by Alzheimer’s disease (AD) (n = 62, 19.1%) and the composite “Other Dementia” category (n = 47, 14.5%; subtypes listed in Fig. 2B legend). PD and the dementia categories combined account for 270 of 325 included studies (83.1%) across the evidence base. The remaining 17% is divided across Mild Cognitive Impairment (MCI; n = 23, 7.1%), Motor Neuron Disease (MND) including Amyotrophic Lateral Sclerosis (ALS) (n = 13, 4.0%), mixed neurodegenerative populations (n = 10, 3.1%), and Huntington’s Disease (n = 9, 2.8%). Each study was assigned to one primary technology domain for analysis, secondary domain assignments are reported in Supplementary Figure 1.

### 2.3 Study Duration and Sample Size

Within this growing evidence base, the studies typically had small sample sizes with short durations. The median sample size was 23 participants (IQR 11–46), and sample sizes ranged from fewer than 10 to more than 2,000 participants (Fig. 3). Evaluation duration was equally limited but split into two qualitatively different design types. Of the 325 studies, 68 (20.9%) used sub-day or single-session designs, comprising single-session usability evaluations, baseline measurements, and other cross-sectional snapshots (41 studies with no adequate duration field in the extraction record, and 27 with quantifiable durations under one day). The remaining 257 studies followed participants for at least a day, with a median duration of 42 days (IQR 7–121) and the longest extending to up to 6 years. As these two groups capture fundamentally different kinds of evaluation, a single interaction with the technology versus sustained observation over time, we treated them separately in all subsequent analyses. The 257 day-or-longer studies form the analytical sample for Figure 3.

**Figure 3.**
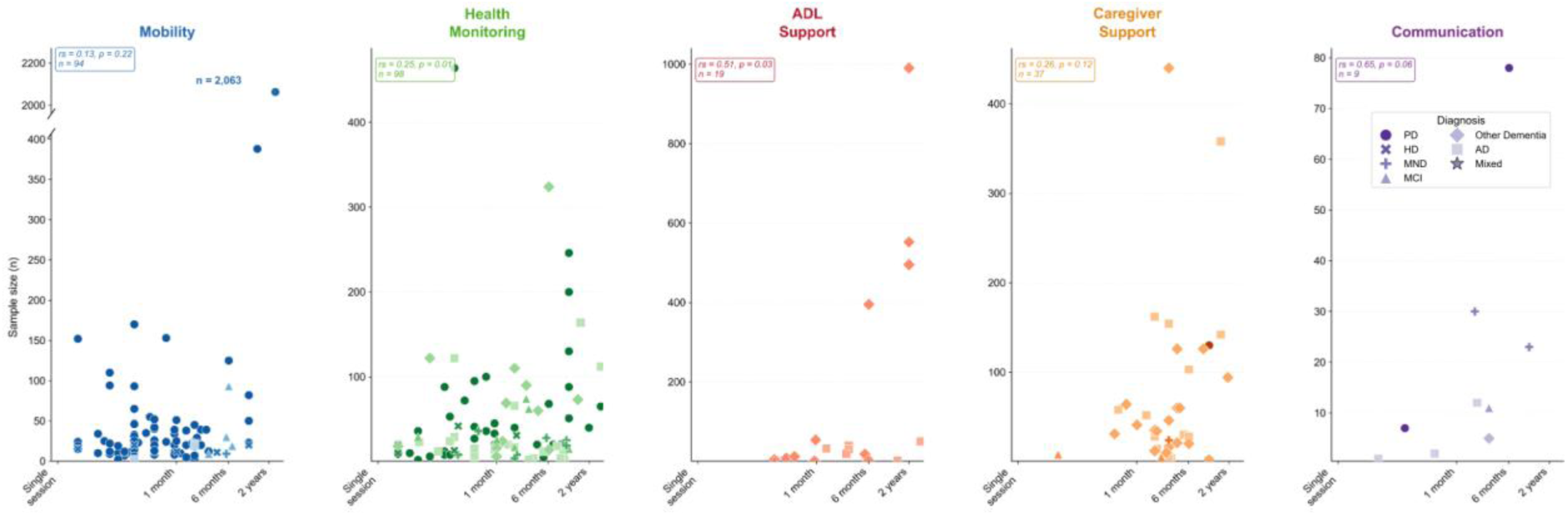
Joint distribution of study duration and sample size across the five technology domains. Scatter plots show sample size (y-axis) against study duration on a log₁₀ scale (x-axis) for each domain. Marker shapes denote primary clinical diagnosis. Spearman rank correlation coefficients (rₛ) and p-values are shown for each panel. The y-axis for Mobility includes a break to accommodate the largest outlier (n = 2,063). Per-panel n values reflect day-or-longer studies with both duration and sample size reported in extractable form; the 68 sub-day or single-session studies are characterised separately in Supplementary Table S5. *Abbreviations*: *AD, Alzheimer’s disease; ADL, activities of daily living; HD, Huntington’s disease; MCI, mild cognitive impairment; MND, motor neuron disease; PD, Parkinson’s disease*.

The joint distribution of duration and sample size varied across domains (Fig. 3). Mobility and Health Monitoring contained the largest number of studies (n = 129 and n = 110, respectively) and the widest spread in both duration and sample size, including the only studies with durations exceeding 6 months and samples exceeding 100 participants. The largest recorded sample (n = 2,063) involved a PD cohort monitored over 2.5 years within the Mobility domain. ADL Support (n = 33) and Caregiver Support (n = 43) contained fewer studies overall but showed a clear tendency for larger samples to accompany longer study durations. Communication was the smallest domain (n = 10) and contained no studies lasting more than 6 months. When stratified by target diagnosis, studies exceeding 6 months in duration that enrolled more than 100 participants were concentrated exclusively within the Mobility, Health Monitoring, and Caregiver Support domains. Within Caregiver Support, longer-duration studies predominantly involved mixed-etiology or unspecified-subtype dementia cohorts rather than single-condition populations. Per-diagnosis study counts, median sample sizes, durations, and percentages of reliance on prototypes are reported in Supplementary Table S1.

Considering the high prevalence of relatively small and short-duration studies, we examined the relationship between sample size and duration. Spearman rank correlations were weak and non-significant in Mobility (rₛ = 0.13, p = 0.22, n = 94) or Caregiver Support (rₛ = 0.26, p = 0.12, n = 37). Modest positive correlations were observed in Health Monitoring (rₛ = 0.25, p = 0.01, n = 98) and ADL Support (rₛ = 0.51, p = 0.03, n = 19). Communication contained too few studies to support inferential interpretation (rₛ = 0.65, p = 0.06, n = 9). In the domains that account for the majority of the literature, sample size and evaluation duration therefore appear to vary largely independently. We identified only 19 studies (5.8% of 325) that were large (N ≥ 50) and long (over 12 months), as presented in Supplementary Table S3.

To summarise overall evaluation effort across all 325 included studies, we calculated participant-days as the product of sample size and observation duration, with sub-day and single-session designs assigned a duration of one day for this calculation. The median study recorded 365 participant-days (IQR 35 to 1,960). The distribution was concentrated at small-effort evaluations: 36% of studies (n = 118) recorded fewer than 100 participant-days, 54% (n = 176) fewer than 500, and 65% (n = 212) fewer than 1,000.

### 2.4 Technology Characteristics, Study Design, and System Origin

The included studies showed convergence in the technologies they used. The technologies clustered around a single configuration. Wearable devices using motion or inertial measurement unit (IMU) sensors (n = 162) were deployed primarily within the Mobility and Health Monitoring domains (Fig. 4A). Other technology and sensor pairings, such as telehealth platforms and mobile applications using input/interaction data (n = 25 and n = 20, respectively), and smart home systems with environmental sensors (n = 14), were considerably less common. The least represented categories were robotics platforms (n = 7) and software-based tools (n = 9), which appeared primarily within the Caregiver Support and ADL Support domains.

**Figure 4.**
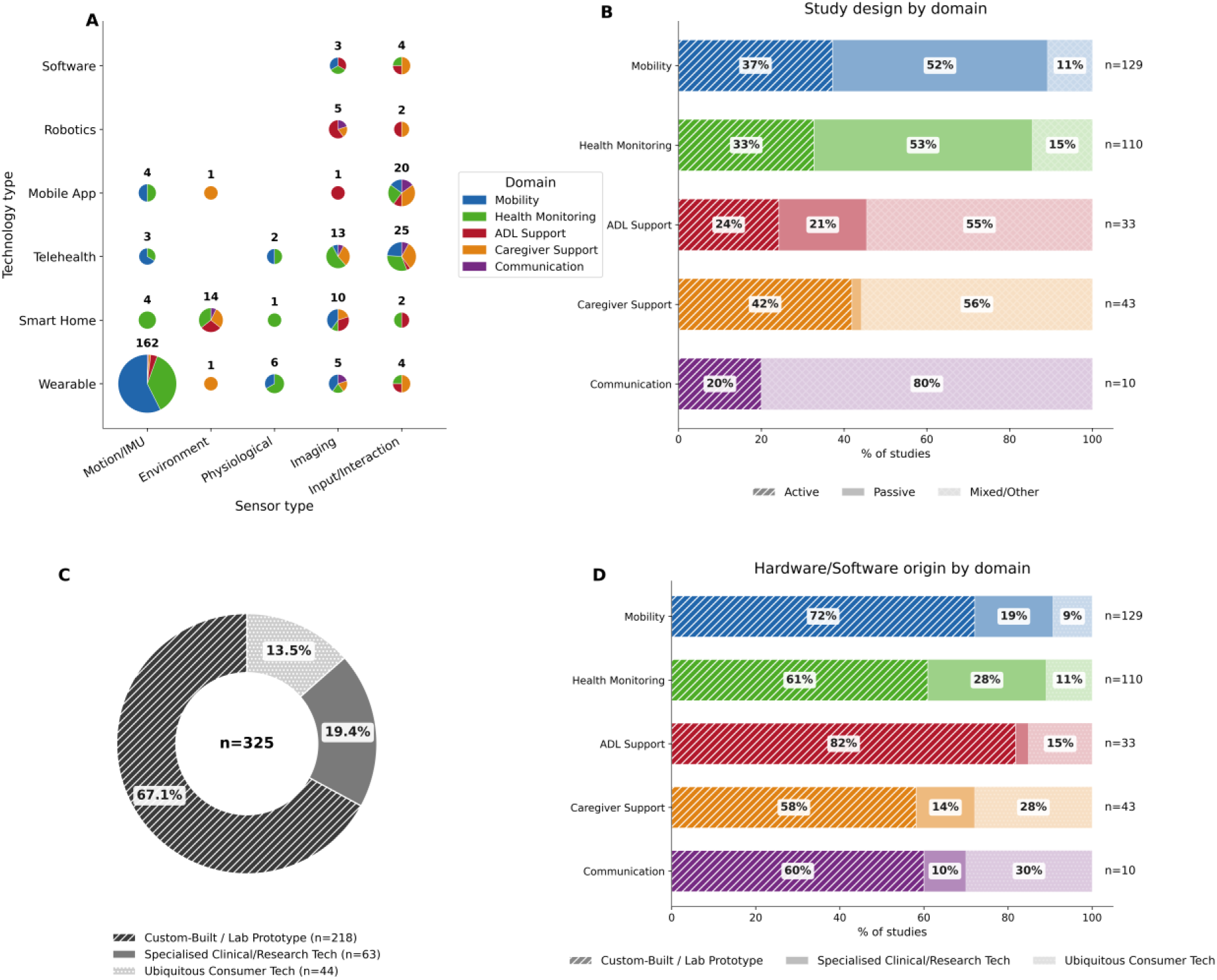
Technology characteristics, study design, and system origin. **(A)** Bubble chart illustrates the frequency of sensor types used within various technology platforms; bubble size corresponds to the number of studies and pie slices indicate the proportion of application domains. **(B)** Distribution of study designs (active, passive, or mixed/other) stratified by application domain. **(C)** Overall proportion of hardware and software origins among the included studies. **(D)** Hardware and software origins categorised by application domain*. Abbreviations: IMU, Inertial Measurement Unit*

This concentration of wearable and motion-based sensing was reflected in how studies were designed (Fig. 4B). Passive, observational designs dominated in Mobility (52%) and Health Monitoring (53%), the same domains where IMU-based wearables were most common. In contrast, ADL Support, Caregiver Support, and Communication relied on mixed or other designs (55%, 56%, and 80% respectively), consistent with their greater use of input/interaction and environmental sensing.

Given these domain-specific patterns in both technology and study design, we examined the technological development of the evidence base to assess whether the field is built primarily on consumer-ready hardware or on research-stage devices (Fig. 4C). Custom-built or lab-prototype technologies dominated (n = 218, 67.1%), while specialised clinical or research-grade technologies accounted for a further 19.4% (n = 63), and ubiquitous consumer technologies represented the smallest segment (n = 44, 13.5%). Combined, 86.5% of included technologies (n = 281) were either custom-built prototypes or specialised research-grade devices. This pattern held across domains (Fig. 4D), with custom-built prototypes most prevalent in ADL Support (82%) and Mobility (72%). Ubiquitous consumer technology was more common in Communication (30%) and Caregiver Support (28%) than in domains focused on physical or medical monitoring.

### 2.5 Usability Evaluation, Adherence, and Acceptability

The maturity of the technology was matched by the maturity of its evaluation. Across the included studies, assessment approaches were characterised by widespread reliance on non-standardised methods, a pattern evident in usability assessment, outcome reporting, and adherence measurement (Fig. 5). Of the 207 studies that assessed usability, 171 (82.6%) relied on custom or unvalidated surveys, while only 32 (15.5%) used standardised validated tools such as the System Usability Scale (SUS), the User Experience Questionnaire (UEQ), or UTAUT, and 4 (1.9%) used clinical scales as proxy measures. This reliance on unvalidated tools was consistent across all domains, ranging from 68% in ADL Support to 86% in Mobility and Health Monitoring (Fig. 5B).

**Figure 5.**
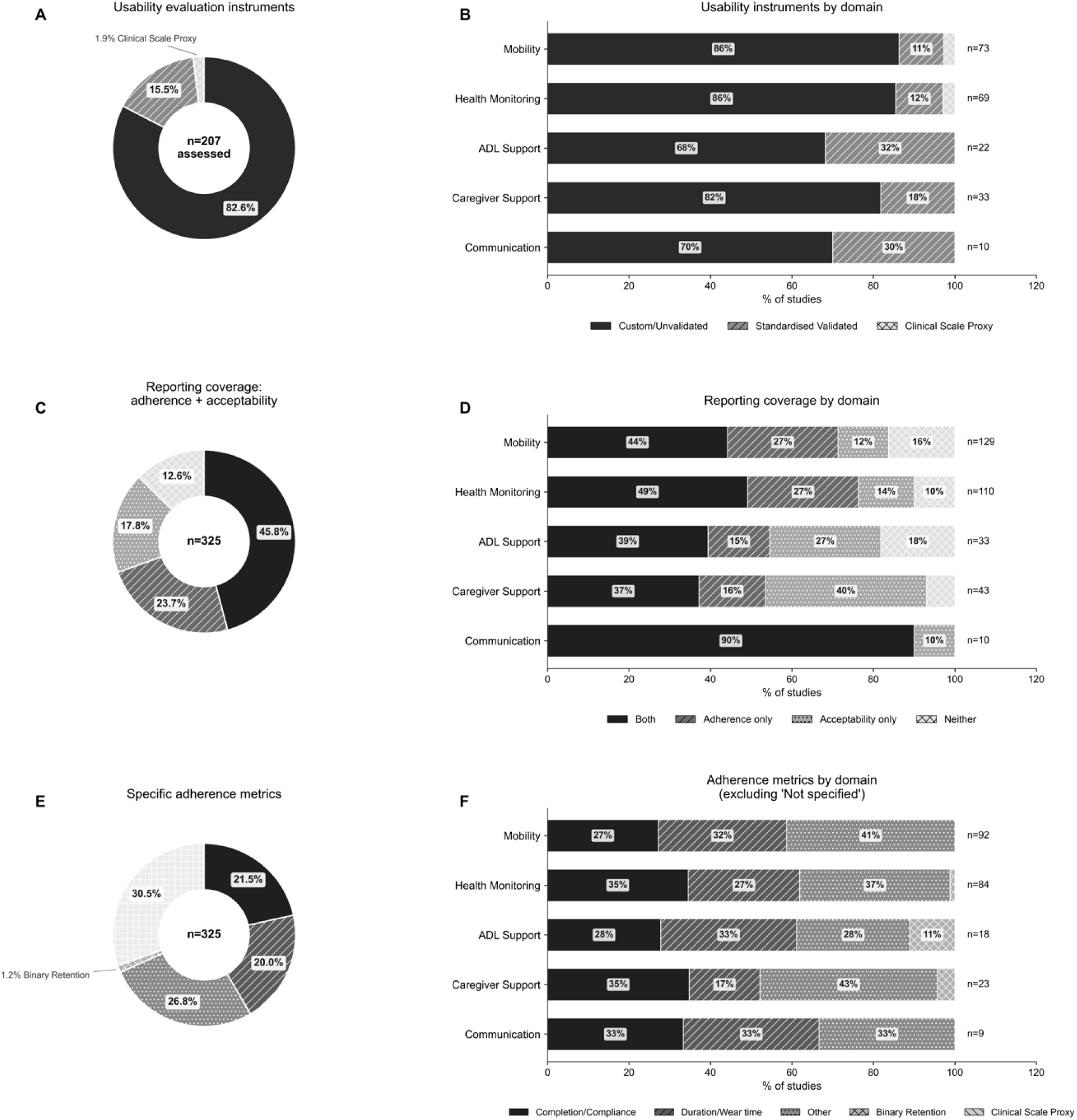
Usability evaluation and adherence reporting across included studies. (A, **B)** Proportion of studies using specific usability evaluation instruments, presented overall **(A)** and stratified by technology domain, with **(B)**. **(C, D)** Reporting coverage of adherence and acceptability outcomes, presented overall **(C)** and across domains **(D)**. **(E, F)** Specific metrics used to quantify adherence, presented overall **(E)** and stratified by domain excluding “Not mentioned” studies **(F).** *Abbreviations: ADL, Activities of Daily Living*.

This broader lack of standardisation extended beyond usability into the reporting of adherence and acceptability outcomes. Both outcomes were reported in 45.8% of studies (n = 149), while 23.7% (n = 77) reported adherence only, 17.8% (n = 58) reported acceptability only, and 12.6% (n = 41) reported neither (Fig. 5C). Combined reporting was highest in Communication (90%, n = 10), Health Monitoring (49%, n = 110), and Mobility (44%, n = 129), whereas absence of both metrics was most common in ADL Support (18%) and Mobility (16%) (Fig. 5D).

Even among studies that reported adherence, the metrics used were often unspecified or non-comparable (Fig. 5E). A total of 30.5% (n = 99) did not specify a clearly defined adherence metric. Where a metric was defined, “Other” metrics that could not be mapped to a standard category were most common overall (26.8%, n = 87), followed by completion or compliance rates (21.5%, n = 70) and duration or wear time (20.0%, n = 65). Domain-level patterns differed, with completion or compliance most common in Health Monitoring and Communication (35% and 33% of those reporting), whereas duration or wear time was more prevalent in domains requiring continuous measurement, particularly ADL Support (33%) and Mobility (32%) (Fig. 5F).

In total, 57.2% of included studies (n = 186) either did not specify an adherence metric or used one that could not be mapped to a standard category. Patterns of reported technical difficulties also varied across platform types and domains, with connectivity/sync (n = 66) and software bugs (n = 65) the most frequently reported issue types and Health Monitoring and Mobility studies disproportionately affected (Supplementary Table S2; Supplementary Figure 2). 155 studies (47.7%) did not report any technical issues.

### 2.6 Barriers and Facilitators

Barriers and facilitators were extracted from all 325 included studies. Each study could contribute to multiple themes if it discussed several types of barriers or facilitators. The totals, therefore, count theme mentions, not studies, and any single study may appear in more than one theme. In total, 763 barrier mentions and 902 facilitator mentions were recorded, with facilitators outnumbering barriers across all domains.

Overall, the reported barriers were predominantly methodological in nature (Fig. 6A). The most frequently reported category was Measurement and Validation (n = 186 mentions), encompassing concerns about sensor accuracy and algorithm validation limitations. This was followed by Environmental and External Factors (n = 160) and Sample Size and Power (n = 92). Study Design Constraints (n = 89), Attrition and Retention (n = 89), and Bias and Generalisability (n = 87) were reported at similar frequencies, while Data Quality and Completeness was the least common (n = 60). The domain distribution of these barriers mirrored the overall volumetric distribution of the literature, with Mobility and Health Monitoring together accounting for most mentions across all seven categories.

**Figure 6.**
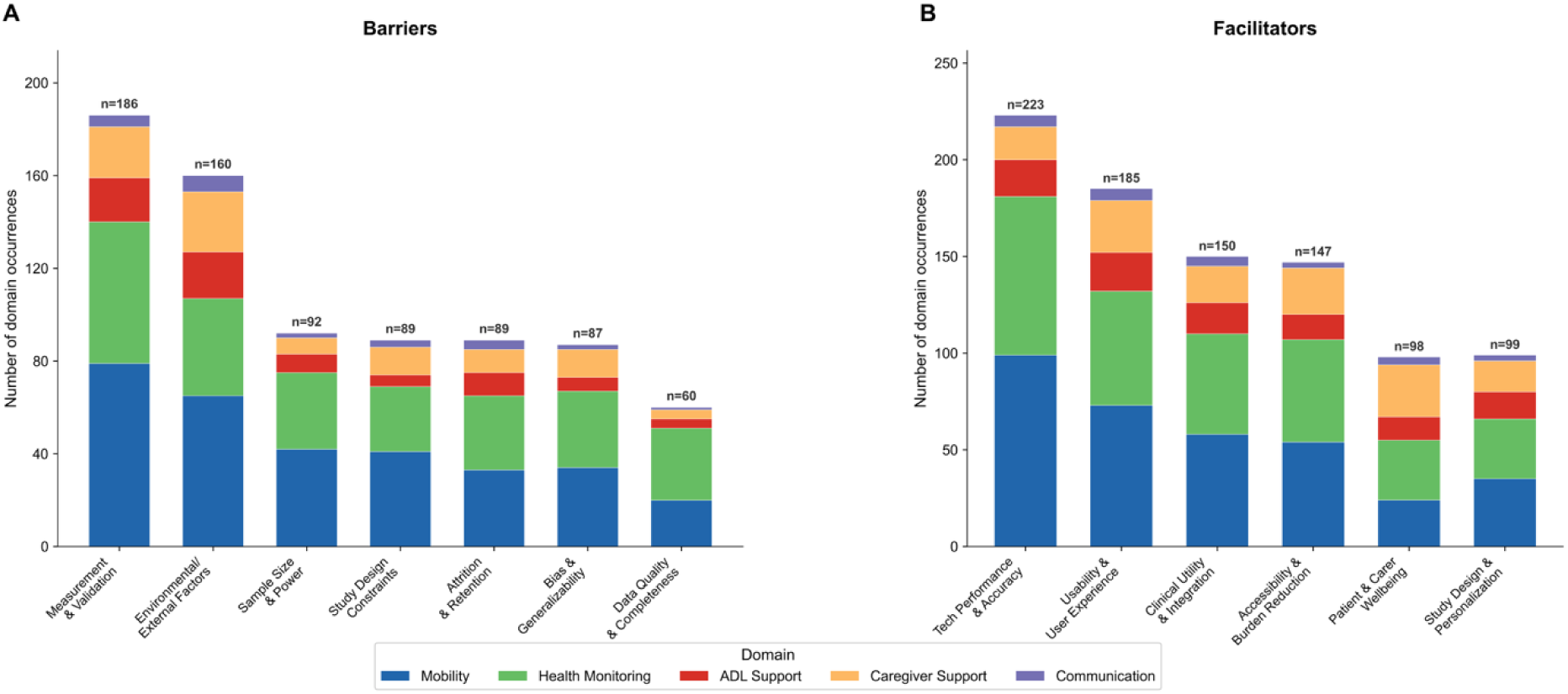
Key barriers and facilitators to technology implementation. **(A)** Frequency of reported barriers, categorised by thematic area and stacked by the primary application domain. **(B)** Frequency of reported facilitators, similarly categorised by theme and stacked by domain. Values represent the total number of study-by-domain occurrences for each theme; individual studies may contribute to multiple themes.

In contrast to these predominantly methodological barriers, facilitators were more commonly framed around the practical and experiential benefits technology delivered to the user (Fig. 6B). Technology Performance and Accuracy was the most frequently cited (n = 223 mentions), followed by Usability, Acceptability and User Experience (n = 185), Clinical Utility and Healthcare Integration (n = 150), and Accessibility, Convenience and Burden Reduction (n = 147). Study Design, Data Quality and Personalisation (n = 99) and Patient and Caregiver Wellbeing (n = 98) were reported at similar frequencies. As with barriers, the absolute frequency of facilitators was highest in Mobility and Health Monitoring, again consistent with the volumetric pattern observed in the literature.

However, a different pattern emerged for the user-centred facilitator categories. Caregiver Support and ADL Support contributed proportionally larger shares of facilitators relating to user experience, wellbeing and accessibility than their overall representation in the literature would predict. Caregiver Support accounted for 27 of 185 usability mentions (14.6%) and 27 of 98 wellbeing mentions (27.6%), despite comprising only 13.2% of included studies.

## Discussion

This scoping review of 325 studies of digital and assistive technologies for neurodegenerative conditions, published between 2001 and 2025, describes a literature growing rapidly in volume but converging slowly on the methodological features and translational pathway needed to support clinical translation. The pattern of small, short, prototype-based evaluations has been described across digital health under the term pilotitis^17,18^. Our findings characterise and quantify this pattern for the neurodegenerative-technology literature specifically and map it against the five technology domains relevant to home-based care.

Digital medicine literature in this field, as captured in this review, reflects a strong bias between domains and disorders. PD accounted for 49.5% of all studies, reflecting the suitability of its motor phenotype for IMU-based measurement compared with other neurodegenerative conditions^22^, while HD and MND remain marginal. Yet, this bias is at least partially driven by different needs, e.g., the MND literature extends into specialist communication and rehabilitation fields^23^ that were not captured by the search strategy. As for domains, Mobility and Health Monitoring, the two domains supported by well-established wearable platforms and sensor pipelines^24^, dominated the evidence base, accounting for 73.5% of all studies. In comparison, Communication and ADL Support have shown comparatively slower growth. The same imbalance appears in the diagnostic distribution. Major areas where technology could have a substantial impact on life quality^7^ and time to institutionalisation^25^ are under-represented in the literature^26^ including technologies for daily function, communication, and caregiver capacity. There is an under-representation of non-motor symptoms^27^, such as cognitive dysfunction^28^, mood disturbances^28^, and fatigue^29^, despite their consistent ranking by patients and caregivers as among the most distressing features of these conditions^30^.

The participant-days distribution reveals relatively few longer-term and/or large-scale studies, with more than a third of studies recording fewer than 100 participant-days, and roughly two-thirds fewer than 1,000. The other features associated with pilotitis are similarly prevalent field-wide, where 67% of the field uses custom-built or prototype hardware, and the lack of usability assessment via standardised, validated tools. The pilotitis pattern described in earlier digital health reviews^17,18^ therefore characterises the current state of the field.

These design features tend to co-occur across individual studies and map directly onto the NASSS framework’s account of why digital health innovations stall during scale-up^31,32^. First, 86.5% of included technologies were custom-built or research-grade hardware, placing most evaluated devices outside the regulatory pathways (FDA, MHRA, CE) required for clinical deployment. Second, the median evaluation window of 14 days is far shorter than the timescales over which falls, cognitive decline, and functional loss progress in these populations, further suggesting that sustained use was rarely observed^33^. Third, 20.3% (66/325) of included studies appeared in engineering journals (e.g., IEEE journals, Sensors, Biomedical Engineering), where prototype validation is both expected and appropriately valued rather than indicative of stalled translation. Excluding these studies, the remaining clinically oriented literature shows a longer median evaluation window (rising from 14 to 28 days) and modestly larger samples, indicating that the translational gap is narrower within work explicitly aimed at clinical deployment. Reliance on custom-built or research-grade hardware nonetheless persists (83.1%), suggesting that limited use of regulatory-ready, deployable technology remains a cross-cutting barrier rather than an artefact of engineering-venue publishing.

Study duration varied by condition but did not consistently track disease progression (Supplementary Table S4). The shortest median durations were observed in PD and HD, the two diagnostic groups most heavily represented by short-window wearable gait monitoring. PD studies were dominated by IMU-based wearable designs running for days rather than weeks, which may be useful for medication titration but ignore the timescales over which falls, changes in gait, and motor impairments typically occur^22,23,34^. Caregiver Support studies ran for months, consistent with the sustained engagement needed to produce measurable outcomes in this area^35^ while most Mobility and ADL studies lasted less than a week, limiting their ability to detect falls and other adverse events, as well as meaningful changes in function and independence^22,23,34^. This highlights the need for funding of longer-term studies.

The relationship between technology availability and study scale was not uniform across domains. In Mobility and Health Monitoring, passive observational designs dominated (52% and 53%, respectively), as IMU based wearables can produce continuous quantitative output without requiring participant action^24,36^. ADL Support, Communication, and Caregiver Support, the three domains without an established sensing approach, relied mainly on mixed or qualitative designs that combined interviews, surveys, and small pilot interventions. The same pattern appeared in the origin of the technologies. In Mobility, Health Monitoring, and ADL Support, the three domains that rely on sensor-based measurement, consumer-grade hardware accounts for less than 16% of included studies. In contrast, Caregiver Support and Communication relied more heavily on telehealth platforms, mobile applications, and consumer communication tools already on the market, with consumer-grade hardware accounting for 28% and 30% of studies in these domains. The participant days distribution mirrors this split, where Caregiver Support and Health Monitoring contribute a larger share of the large-scale studies than Mobility, despite Mobility being the most populous domain. As expected, considering the complexity and likely cost of developing appropriate and safe ADL Support, this domain had the highest level of prototype hardware, small samples and short duration studies. This is despite ADL impairment being among the strongest predictors of admission to residential care^37^, hence demonstrating a clear evidence gap in a key area with substantial economic and quality-of-life implications, consistent with recent literature identifying limited evidence for ADL-related technologies more broadly^16^.

Among studies that assessed usability (63.7%), only 15.5% (32 of 207) used standardised tools such as the System Usability Scale (SUS), the User Experience Questionnaire (UEQ), or UTAUT (Fig. 5A), while the remainder relied on custom or unvalidated instruments. When each study uses its own usability survey, findings cannot be meaningfully compared across papers, limiting opportunities for quantitative synthesis. In addition, standardised tools raise questions about population-specific validity. SUS was developed and normed in cognitively intact adults, and its validity in populations with moderate AD or motor and speech impairments has not been comprehensively established^38^. Adherence reporting showed similar inconsistencies. 57.2% of studies reporting adherence either omitted a metric or used one that could not be mapped to a standard category. Domains also differed in how adherence was measured. Completion-based measures dominated in Communication and Health Monitoring, whereas wear-time measures were more common in Mobility.

A structured taxonomy and associated core measurement strategy, considering adoption, adherence, persistence, and optimal use as distinct constructs, could improve comparability across studies and research efficacy^38^. These observations are consistent with the 2025 INTERDEM AI and Assistive Technology Taskforce review of 112 systematic reviews^39^, which also called for standardised and theory-informed evaluation methods. Most of the included literature sits at the development and feasibility phases of the Medical Research Council (MRC) framework^20^, with comparatively few reaching the scale typical of its evaluation phase. As scoping reviews cannot determine whether individual studies progressed to evaluation or implementation outside of our search dates, it is important to note that our framework characterises the structure of the published field as a whole, rather than the adequacy of any individual study. However, future studies integrating usability, adherence, and outcome evaluation within a single development pathway, rather than treating each as a separate decision, would help build cumulative evidence that the framework is designed to produce.

As a scoping review, this synthesis describes the literature in aggregate rather than the appropriateness of any individual study, and the English-language restriction may have excluded relevant work from settings where digital health research is active. The OSF protocol was registered after the database search and during data extraction, so registration was retrospective rather than fully prospective; the registered protocol covers the eligibility criteria, search strategy, and primary mapping aims, while the participant-days analysis was developed post hoc during synthesis.

The findings from this review describe a literature that has expanded in volume while slowly converging on the methodological features needed to produce cumulative and comparable evidence. Most studies remain small and short, built on prototype hardware and evaluated with custom instruments; only a small minority combine larger cohorts, longer follow-up, established platforms, and validated tools. The pattern is consistent with a field still operating predominantly at the feasibility stage of the MRC development pathway^40^. The clinical priorities associated with neurodegenerative conditions, including progressive loss of mobility, function, communication, and caregiver capacity^27,41^, have changed little over the past two decades. However, the rate at which feasibility stage evidence is being published now appears to exceed the rate at which translationally meaningful evidence is being produced. A greater convergence around shared platforms, evaluation methods, and validation pathways aligned with disease timescales is needed, in order to translate this research field into real-world impact and meaningful gains in clinical utility.

## Methods

### Study Design

This scoping review was conducted in accordance with the Joanna Briggs Institute (JBI) methodological framework^42^ for scoping reviews and reported following the PRISMA Extension for Scoping Reviews (PRISMA-ScR)^43^ (Supplementary Material 1). The JBI framework informed the development of the research question, eligibility criteria, search strategy, data extraction, and synthesis processes. This review protocol was registered on the Open Science Framework on December 1^st^ 2025 (OSF; https://doi.org/10.17605/OSF.IO/QJZC9)^44^. Registration was retrospective, occurring after the database search (conducted November 5^th^ 2025) and during data extraction. Registered protocol reflects the eligibility criteria, search strategy, and primary mapping aims of the review. The participant-days summary reported at the end of Section 2.3 was not prespecified. It was developed during synthesis to summarise patterns observed in the extracted data and is a post-hoc analytical contribution.

### Research Question

This review addresses the overarching question: what is the nature of evidence regarding digital and assistive technologies designed to support people with dementia and related neurodegenerative conditions. Our question is structured around four sub questions: (1) what technologies currently exist for this population, in relation to their type, sensor modality, and system origin; (2) how do these technologies map across the five domains (Mobility, Health Monitoring, Activities of Daily Living, Caregiver Support, Communication); (3) how have studies evaluated usability, acceptability, and adherence; and (4) what study level characteristics, such as study design, sample size, duration, and reported barriers and facilitators, are reported across the evidence base?

### Search Strategy

The search strategy was developed collaboratively by the review team with input from a subject librarian, using the Population, Concept, Context (PCC) framework^42^ and the five technology domains defined by the ZeDTech Network (Mobility, Communication, Activities of Daily Living, Health Monitoring, Caregiver Support; Table 1 and Supplementary Material 2) to guide keyword development. A preliminary scoping search was conducted in MEDLINE and CINAHL in October 2025 to test sensitivity and refine the search string. Eight electronic databases were then searched systematically from inception to 5 November 2025: IEEE Xplore, CINAHL, Embase, Scopus, MEDLINE, PsycINFO, Global Health, and AMED. The search strategy was structured around four main blocks, Population, Technology, Domains, and Context, with Medical Subject Headings (MeSH) terms and free-text keywords combined using ‘OR’ within each block and ‘AND’ across blocks for the final search set. Proximity operators were applied where supported, and the strategy was adapted to the indexing conventions of each platform. To maintain a defined and reproducible search frame within available resources, the search was restricted to peer-reviewed literature in English; grey literature, conference proceedings beyond those indexed in IEEE Xplore, and forward and backward citation searching were not undertaken. Full search string is provided in Supplementary Material 3.

**Table 1.** Definitions of ZeDTech technology domains & Operational Definitions.

| Technology Domains |  |  |
| --- | --- | --- |
| Technology Domain | Definitions |  |
| Mobility | Technologies that assess, monitor or support movement, gait, balance, and physical activity to promote safe and independent mobility |  |
| Communication | Technologies that facilitate interaction between individuals, caregivers, and clinicians, including speech, messaging and assistive communication interfaces |  |
| Activities of Daily Living | Technologies that support activities of daily living, enabling independence in tasks such as dressing, eating, hygiene, and home functioning |  |
| Health Monitoring | Technologies that capture, track and analyse physiological, behavioural or clinical data to support ongoing health assessment and management |  |
| Caregiver Support | Technologies designed to support caregivers through monitoring, coordination, education and emotional support or social support, reducing burden and enhancing care delivery |  |
| Operational Definitions |  |  |
| Operational Terms | Definitions |  |
| Adherence | The extent to which user engagement with the technology aligns with the prescribed or intended usage protocol over the study period |  |
| Usability | The extent to which a system enables users to achieve specified goals with effectiveness, efficiency and satisfaction in a defined context of use |  |
| Feasibility | The extent to which a technology can be successfully utilised as intended, including recruitment, retention, acceptability and practicality within the study context |  |
| Implementation Facilitators and Barriers | Factors that enable or hinder the adoption, integration and sustained use of a technology within real-world settings, including individual, technological and system-level influences |  |
| Operational Definitions for Hardware/Software Origin |  |  |
| Category | Definition | Examples |
| Custom-Built / Lab Prototype | Hardware built or substantially modified by the research team; algorithms or systems with no commercial or research-grade equivalent. | Research-team-built PCB wearable; in-house smartphone application without public release. |
| Specialised Clinical / Research Tech | Commercially marketed products designed for research or clinical users, deployed as the manufacturer intended. | ActiGraph, GENEActiv, Axivity, Empatica E4, PDMonitor, Personal KinetiGraph, Tobii eye-trackers. |
| Ubiquitous Consumer Tech | Mass-market products sold to the general public and used essentially as shipped. | Fitbit, Apple Watch, Withings, Oura, Zoom, FaceTime. |

### Eligibility criteria

**Participants:** Studies were eligible if they included adults (≥18 years) with a clinical diagnosis of dementia of any type (including AD, VD, frontotemporal dementia, dementia with Lewy bodies, Parkinson’s disease dementia, mixed dementia, and unspecified dementia), MCI, PD, MND, ALS or HD, regardless of sex, disease duration, or severity. Studies involving formal or informal caregivers (e.g., professional care workers, healthcare providers, family members, spouses, and unpaid caregivers) of people with the above conditions were included. Studies with mixed populations (e.g., a combination of included neurodegenerative conditions, or one included condition combined with cognitively healthy older adults) were included if data for the relevant included population subgroup were reported separately. Studies in which dementia, MCI, PD, MND, or HD was not the primary condition of interest, studies where cognitive impairment was a secondary outcome, and studies involving only animal subjects were excluded. For analysis and figure labelling, ALS was grouped under MND, reflecting that ALS is the most common form of MND and that the two terms are often used interchangeably in the literature. In addition, AD was reported as a distinct category given its volume in the literature, while studies targeting non-AD, mixed-etiology dementia, and unspecified-subtype dementia were grouped as “Other Dementia”.

**Concept:** Eligible studies investigated technology-based interventions or monitoring systems used in home or community settings, including wearable devices (e.g., smartwatches, activity trackers), remote monitoring systems (e.g., smart home sensors, ambient sensors), or assistive technologies (e.g., GPS devices, medication reminders, social robots, mobile applications, voice assistants). Studies addressing at least one of the following predefined technology domains: (1) mobility monitoring or support (e.g., fall detection, wandering prevention, gait monitoring); (2) ADL support (e.g., medication adherence, self-care assistance); (3) health monitoring (e.g., vital signs, sleep, behavioural symptoms); (4) caregiver support (e.g., burden reduction, respite care, training); or (5) communication support (e.g., social engagement, cognitive stimulation, reminders). Studies that do not include a technology-based intervention, monitoring system, or assistive device (e.g., purely pharmacological or non-technological behavioural interventions), and studies that do not address any of the five technology domains.

**Context:** Studies conducted in home-based, community, independent living, or laboratory settings were eligible. Laboratory and simulated-home studies were included to capture the full spectrum from proof-of-concept to real-world deployment, as excluding early-stage evaluations would obscure the maturity profile of the evidence base. Studies conducted in mixed settings (e.g., hospital and home) were included where home- or community-based outcomes were reported separately. Studies conducted exclusively in institutional settings (e.g., acute hospitals, nursing homes, long-term care facilities, or residential aged care facilities) without any home- or community-based component, or without separating reporting of such outcomes were excluded.

**Study designs:** Eligible designs included randomised and non-randomised controlled trials, before-and-after studies, interrupted time-series studies, cohort studies, case-control studies, analytical and descriptive cross-sectional studies, case series and case reports, qualitative studies (including phenomenology, grounded theory, ethnography, qualitative description, action research, and feminist research), and mixed-methods studies. Only studies published in English were included, owing to the absence of translation resources within the review team. Reviews of any type, conference abstracts, dissertations, book chapters, editorials, commentaries, letters, study protocols without results, other grey literature, and studies for which full text was not accessible were excluded.

**Screening and study selection:** All records identified through database searching were imported into Covidence (Veritas Health Innovation, Melbourne, Australia) for deduplication and review management. A two-stage screening process was conducted. Titles and abstracts were independently screened against the eligibility criteria by two reviewers (SB and MF). Full texts of potentially eligible records were then sought through institutional library access. If full texts could not be retrieved, the records were excluded. Retrieved full texts were independently assessed for inclusion by the same two reviewers, with inter-rater agreement at this stage of κ = 0.97. Disagreements at each stage were resolved through discussion, with a third reviewer (SC), where consensus could not be reached. The screening process and reasons for exclusion at the full-text stage are presented in the PRISMA-ScR flow diagram (Fig. 1).

**Data extraction:** Data were extracted into a standardised spreadsheet using a pre-tested extraction instrument developed in accordance with the JBI Manual for Evidence Synthesis^45^. Data extraction was led by SB, with a random 25% subset extracted by MF and a further 2% subset extracted by SC. Verification was based on exact concordance between independent category assignments before any reconciling discussion. Where discrepancies arose, they were resolved through team discussion.

**Data synthesis:** Given the heterogeneity of included studies, a narrative synthesis approach was adopted, consistent with the descriptive and mapping aims of a scoping review. Extracted data were coded by SB using a hybrid deductive-inductive approach. A preliminary deductive framework based on the predefined technology domains and study characteristics was expanded with inductive codes to capture qualitative themes including implementation barriers and facilitators, sensor types, and technology categories (Table 2). The coding scheme was piloted on 20 studies, refined through team discussion, and then applied consistently across all included studies. Uncertainties in category assignment were resolved by team consensus. Coded data were synthesised descriptively and presented in tabular and graphical form to illustrate the distribution of study characteristics across the five technology domains. Where technologies spanned multiple functional areas, a primary domain was assigned based on principal intended function; secondary domains were recorded separately. Primary domain assignment was made by SB and reviewed by MF, with discrepancies resolved through discussion. Descriptive statistics (frequencies, proportions, medians) were calculated for sample size, study duration, and hardware origin. To summarise overall evaluation effort, participant-days (sample size × observation duration) were computed for each study, with sub-day and single-session designs assigned a duration of one day for this calculation. The resulting distribution was characterised using percentile thresholds and median (with IQR) rather than mean values given the wide range of the data. These analyses are exploratory; the included studies represent the accessible published literature rather than a probability sample, and results should be interpreted as characterising observed patterns rather than estimating population parameters. All analyses were conducted in Python (3.12.7) using pandas (2.1.3), SciPy (1.13.1), and Matplotlib (3.8.2). Extracted data and analysis code are available as detailed in the Data and Code Availability statements.

**Table 2.** Key information and inductive coding categories for data extraction.

| **Grouping Category** | **Categories / Coding Scheme** |
| --- | --- |
| **Country of Origin** | Country of study publication; multi-country studies classified by majority vote (tie resolved as HIC); income classification per World Bank criteria (HIC / LMIC) |
| **Publication Year Range** | Year of publication; used to characterise temporal trends across the review period (2001–2025) |
| **Study Design** | Passive (analytical observational, descriptive) / Active (interventional, RCT, non-RCT) / Mixed Methods / Qualitative |
| **Technology Type** | Wearable / Smart Home / Telehealth / Mobile Application / Software / Robotics |
| **Sensor Type** | Motion/IMU (accelerometer, gyroscope, magnetometer, actigraph) / Physiological (ECG, PPG, pulse oximetry, blood pressure) / Imaging (camera, video, eye-tracker) / Environment (PIR, contact sensor, smart plug, temperature) / Input/Interaction (touchscreen, microphone, GPS, EEG) |
| **Implementation Stage** | Custom-Built prototype / Specialised research-grade / Consumer Technology (commercially marketed) |
| **Population** | Primary diagnosis (PD / AD / Other Dementia / MCI / ALS/MND / HD); participant type (patient / caregiver / mixed) |
| **Caregiver Type** | Informal (family member, spouse, unpaid carer) / Paid/Professional (nurse, care manager, care worker) / Not specified |
| **Technology Domain (Primary)** | Mobility / Health Monitoring / ADL Support / Caregiver Support / Communication |
| **Study Duration** | Extracted in original units and converted to days for analysis; categorised as: single session (<1 day) / short (<30 days) / medium (30–364 days) / long (≥365 days) |
| **Sample Size** | Total participant count (cognitive disorder group + motor disorder group where both reported); used to derive domain-level medians and threshold analyses ($n \geq 50$; $n \geq 200$) |
| **Usability Assessment** | Tool type: Standardised validated (SUS, UES, TAM, UEQ, UTAUT, Almere Model) / Custom/Unvalidated survey / Clinical scale proxy / Not evaluated |
| **Adherence Metric** | Presence/absence; metric type: wear time / session completion / % protocol completed / days of valid data / qualitative/binary / custom metric / not reported |
| **Technology Issues** | Presence/absence of reported technology problems; type (connectivity, battery, data loss, hardware) |
| **Key Barriers** | Measurement and Validation / Environmental and External Factors / Sample Size and Power / Study Design Constraints / Attrition and Retention / Bias and Generalisability |
| **Key Facilitators** | Tech Performance and Accuracy / Usability and User Experience / Clinical Utility and Healthcare Integration / Accessibility and Burden Reduction / Patient and Carer Wellbeing / Study Design and Data Quality |

**Definitions of key classifications.** Technologies were assigned to one of three mutually exclusive origin categories based on the primary contribution evaluated in each study. Custom-Built / Lab Prototype denotes technologies developed by the research team that did not exist prior to the study, including bespoke hardware, novel algorithms, and research-developed platforms. Specialised Clinical/Research Tech denotes commercially marketed products designed for research or clinical users and deployed as the manufacturer intended (e.g., ActiGraph, GENEActiv, Axivity, Empatica E4, PDMonitor, Personal KinetiGraph, Tobii eye-trackers), and not available through general consumer channels. Ubiquitous Consumer Tech denotes mass-market products sold to the public and used essentially as shipped (e.g., Fitbit, Apple Watch, Withings, Oura, consumer videoconferencing platforms). Borderline classifications were resolved through team discussion using the operational rules in Table 1.

Study design was classified into three categories. Active studies were those in which the technology functioned as an intervention to change a clinical, behavioural, or functional outcome, including randomised and non-randomised controlled trials and before-and-after evaluations. Passive studies were those in which the technology was used observationally to capture or characterise participant data without acting as an intervening factor, including analytical observational designs and sensor-based monitoring. Mixed/Other captured qualitative studies, mixed methods design, and studies in which the technology served both observational and interventional functions.

## Data availability

The data supporting the findings of this review are available within the article and its supplementary materials. Extracted information from the included studies is summarised in the text and tables.

## Acknowledgments

This work was supported by the EPSRC-NIHR Network Plus, ZedTech (grant no. UKRI687). We thank all members of the ZedTech Network for their contribution to the discussions on this review.

HD and CF are supported by the National Institute for Health and Care Research (NIHR) HealthTech Research Centre in sustainable innovation and the Exeter Biomedical Research Centre. The views expressed are those of the author(s) and not necessarily those of the NIHR or the Department of Health and Social Care.

## Author contributions

HD and SH conceptualised the study. SB, MF, SC, JJ, HD and SH developed the review protocol. SB, MF and SC conducted screening and extracted the data. SB, MF, SC, HD and SH conducted analyses. SB and MF drafted the manuscript. All authors reviewed and revised the manuscript.

## References

1 Erkkinen, M. G., Kim, M. O. & Geschwind, M. D. Clinical Neurology and Epidemiology of the Major Neurodegenerative Diseases. Cold Spring Harb. Perspect Biol. 10, a033118 (2018). 10.1101/cshperspect.a033118

2 Przedborski, S., Vila, M. & Jackson-Lewis, V. Series Introduction: Neurodegeneration: What is it and where are we? J. Clin. Invest. 111, 3–10 (2003). 10.1172/JCI17522

3. World Health Organization. *Dementia*, <https://www.who.int/news-room/fact-sheets/detail/dementia> (2025).

4. 4 Alzheimer’s Disease International. *Dementia statistics*, <https://www.alzint.org/about/dementia-facts-figures/dementia-statistics/> (2026).

5 Cummings, J. et al. Alzheimer’s disease drug development pipeline: 2024. Alzheimers Dement. (N. Y.) 10, e12465 (2024). 10.1002/trc2.12465

6 Smith, B. & Ownby, R. L. Disease-Modifying Treatments and Their Future in Alzheimer’s Disease Management. Cureus 16, e56105 (2024). 10.7759/cureus.56105

7 De Marchi, F. et al. Telehealth in Neurodegenerative Diseases: Opportunities and Challenges for Patients and Physicians. Brain Sci. 11, 237 (2021). 10.3390/brainsci11020237

8 Office for Health Improvement and Disparities. *Dementia profile: prevalence and supporting well topics statistical commentary**, March* 2025 https://www.gov.uk/government/statistics/dementia-profile-march-2025-update/dementia-profile-prevalence-and-supporting-well-topics-statistical-commentary-march-2025 (2025).

9 Gamble, L. D. et al. Prevalence of living alone with dementia and other progressive neurological conditions: findings from primary care data in England. BMC Med. 23, 607 (2025). 10.1186/s12916-025-04443-x

10 Parekh, A. K., Goodman, R. A., Gordon, C. & Koh, H. K. Managing multiple chronic conditions: a strategic framework for improving health outcomes and quality of life. Public Health Rep. 126, 460–471 (2011). 10.1177/003335491112600403

11. 11 National Academies of Sciences, Engineering, and Medicine. Social Isolation and Loneliness in Older Adults: Opportunities for the Health Care System (2020). 10.17226/25663

12 Voss, S. et al. Home or hospital for people with dementia and one or more other multimorbidities: What is the potential to reduce avoidable emergency admissions? The HOMEWARD Project Protocol. BMJ Open 7, e016651 (2017). 10.1136/bmjopen-2017-016651

13 Ameri, F. et al. Exploring Caregiver Burden in Alzheimer’s Disease: The Predictive Role Of Psychological Distress. Open Public Health J. 17 (2024). 10.2174/0118749445327572240916091208

14 Ditton, A., Alodan, H., Fox, C., Evans, S. & Cross, J. Exploring the effectiveness and experiences of people living with dementia interacting with digital interventions: A mixed methods systematic review. Dementia 24, 506–551 (2025). 10.1177/14713012241302371

15 Wittich, L. et al. Navigating the complexities of digital health technology implementation: a scoping review of barriers and facilitators. *Implement*. Sci. Commun. 7, 69 (2026). 10.1186/s43058-026-00892-4

16 Jeyasingh-Jacob, J. et al. Markerless Motion Capture to Quantify Functional Performance in Neurodegeneration: Systematic Review. JMIR Aging 7, e52582 (2024). 10.2196/52582

17 Huang, F., Blaschke, S. & Lucas, H. Beyond pilotitis: taking digital health interventions to the national level in China and Uganda. *Glob*. Health 13, 49 (2017). 10.1186/s12992-017-0275-z

18 Egermark, M., Blasiak, A., Remus, A., Sapanel, Y. & Ho, D. Overcoming Pilotitis in Digital Medicine at the Intersection of Data, Clinical Evidence, and Adoption. Adv. Intell. Syst. 4, 2200056 (2022). 10.1002/aisy.202200056

19 Greenhalgh, T. et al. Beyond Adoption: A New Framework for Theorizing and Evaluating Nonadoption, Abandonment, and Challenges to the Scale-Up, Spread, and Sustainability of Health and Care Technologies. J. Med. Internet Res. 19, e367 (2017). 10.2196/jmir.8775

20 Neal, D. et al. Digital assistive technologies for community-dwelling people with dementia: A systematic review of systematic reviews by the INTERDEM AI & assistive technology taskforce. *Digit*. Health 11, 20552076251362353 (2025). 10.1177/20552076251362353

21 Boyle, L. D., Husebo, B. S. & Vislapuu, M. Promotors and barriers to the implementation and adoption of assistive technology and telecare for people with dementia and their caregivers: a systematic review of the literature. BMC Health Serv. Res. 22, 1573 (2022). 10.1186/s12913-022-08968-2

22 Lord, S. et al. Predicting first fall in newly diagnosed Parkinson’s disease: Insights from a fall-naïve cohort. Mov. Disord. 31, 1829–1836 (2016). 10.1002/mds.26742

23 Hunter, H., Rochester, L., Morris, R. & Lord, S. Longitudinal falls data in Parkinson’s disease: feasibility of fall diaries and effect of attrition. Disabil. Rehabil. 40, 2236–2241 (2018). 10.1080/09638288.2017.1329357

24 Zhao, H. et al. Wearable sensors and features for diagnosis of neurodegenerative diseases: A systematic review. *Digit*. Health 9, 20552076231173569 (2023). 10.1177/20552076231173569

25 Bossen, A. L., Kim, H., Williams, K. N., Steinhoff, A. E. & Strieker, M. Emerging roles for telemedicine and smart technologies in dementia care. Smart Homecare Technol. Telehealth 3, 49–57 (2015). 10.2147/SHTT.S59500

26 Evans, J., Brown, M., Coughlan, T., Lawson, G. & Craven, M. P. in Human-Computer Interaction: Interaction Technologies (ed. Kurosu, M.) 406–417 (Springer, 2015). 10.1007/978-3-319-20916-6_38

27 Tosin, M. H., Goetz, C. G. & Stebbins, G. T. Patient With Parkinson Disease and Care Partner Perceptions of Key Domains Affecting Health-Related Quality of Life. Neurology 102, e208028 (2024). 10.1212/WNL.0000000000208028

28 Bock, M. A., Brown, E. G., Zhang, L. & Tanner, C. Association of Motor and Nonmotor Symptoms With Health-Related Quality of Life in a Large Online Cohort of People With Parkinson Disease. Neurology 98, e2194–e2203 (2022). 10.1212/WNL.0000000000200113

29 Pitton Rissardo, J., et al. Exploring Fatigue in Parkinson’s Disease: A Comprehensive Literature Review. Cureus 17, e81129 (2025). 10.7759/cureus.81129

30 van Wamelen, D. J. et al. Digital health technology for non-motor symptoms in people with Parkinson’s disease: Futile or future? Parkinsonism Relat. Disord. 89, 186–194 (2021). 10.1016/j.parkreldis.2021.07.032

31 Greenhalgh, T. et al. Analysing the role of complexity in explaining the fortunes of technology programmes: empirical application of the NASSS framework. BMC Med. 16, 66 (2018). 10.1186/s12916-018-1050-6

32 Guu, T.-W. et al. Wearable devices: underrepresentation in the ageing society. *Lancet Digit*. Health 5, e336–e337 (2023). 10.1016/S2589-7500(23)00069-9

33 Cejudo, A., Arrojo, M., Martín, C. & Almeida, A. AI and Wearables for Early Detection of Cognitive Impairment and Dementia: Systematic Review. J. Med. Internet Res. 28, e86262 (2026). 10.2196/86262

34 Allen, N. E., Schwarzel, A. K. & Canning, C. G. Recurrent Falls in Parkinson’s Disease: A Systematic Review. Parkinsons Dis. 2013, 906274 (2013). 10.1155/2013/906274

35 Liang, M., Rong, J., Grosan, C. & Wang, X. Effectiveness of eHealth Interventions in Alleviating Burden on Informal Caregivers of People With Dementia: Systematic Review and Meta-Analysis of Randomized Controlled Trials. J. Med. Internet Res. 28, e78568 (2026). 10.2196/78568

36 Sapienza, S. et al. Assessing the clinical utility of inertial sensors for home monitoring in Parkinson’s disease: a comprehensive review. npj Parkinsons Dis. 10, 161 (2024). 10.1038/s41531-024-00755-6

37 Gaugler, J. E., Duval, S., Anderson, K. A. & Kane, R. L. Predicting nursing home admission in the U.S: a meta-analysis. BMC Geriatr. 7, 13 (2007). 10.1186/1471-2318-7-13

38 Holden, R. A Simplified System Usability Scale (SUS) for Cognitively Impaired and Older Adults. Proc. Int. Symp. Hum. Factors Ergon. Health Care 9, 180–182 (2020). 10.1177/2327857920091021

39 Sieverink, F., Kelders, S. M. & van Gemert-Pijnen, J. E. Clarifying the Concept of Adherence to eHealth Technology: Systematic Review on When Usage Becomes Adherence. J. Med. Internet Res. 19, e402 (2017). 10.2196/jmir.8578

40 Skivington, K. et al. A new framework for developing and evaluating complex interventions: update of Medical Research Council guidance. BMJ 374, n2061 (2021). 10.1136/bmj.n2061

41 Politis, M. et al. Parkinson’s disease symptoms: The patient’s perspective. Mov. Disord. 25, 1646–1651 (2010). 10.1002/mds.23135

42 Peters, M. D. J. et al. Updated methodological guidance for the conduct of scoping reviews. JBI Evid. Synth. 18, 2119–2126 (2020). 10.11124/jbies-20-00167

43 Tricco, A. C. et al. PRISMA Extension for Scoping Reviews (PRISMA-ScR): Checklist and Explanation. Ann. Intern. Med. 169, 467–473 (2018). 10.7326/m18-0850

44 Birgen, S., et al. Technologies Supporting People with Dementia in Home and Community Settings: A Scoping Review Protocol. (OSF, 2025).

45 Aromataris, E., Lockwood, C., Porritt, K., Pilla, B. & Jordan, Z. JBI Manual for Evidence Synthesis. (JBI, 2024).

