## Supplementary Materials for "Mapping Digital and Assistive Technologies for Dementia and Related Neurodegenerative Conditions: A Scoping Review"

### Supplementary Material 1: PRIMSA checklist

| SECTION | ITEM | PRISMA-ScR CHECKLIST ITEM | REPORTED ON PAGE # |
|---|---|---|---|
| **TITLE** | | | |
| Title | 1 | Identify the report as a scoping review. | 1 |
| **ABSTRACT** | | | |
| Structured summary | 2 | Provide a structured summary that includes (as applicable): background, objectives, eligibility criteria, sources of evidence, charting methods, results, and conclusions that relate to the review questions and objectives. | 1 |
| **INTRODUCTION** | | | |
| Rationale | 3 | Describe the rationale for the review in the context of what is already known. Explain why the review questions/objectives lend themselves to a scoping review approach. | 2-3 |
| Objectives | 4 | Provide an explicit statement of the questions and objectives being addressed with reference to their key elements (e.g., population or participants, concepts, and context) or other relevant key elements used to conceptualize the review questions and/or objectives. | 15 |
| **METHODS** | | | |
| Protocol and registration | 5 | Indicate whether a review protocol exists; state if and where it can be accessed (e.g., a Web address); and if available, provide registration information, including the registration number. | 15 |
| Eligibility criteria | 6 | Specify characteristics of the sources of evidence used as eligibility criteria (e.g., years considered, language, and publication status), and provide a rationale. | 16 |
| Information sources* | 7 | Describe all information sources in the search (e.g., databases with dates of coverage and contact with authors to identify additional sources), as well as the date the most recent search was executed. | 15 |
| Search | 8 | Present the full electronic search strategy for at least 1 database, including any limits used, such that it could be repeated. | Supplementary Material3 (P.4-8) |
| Selection of sources of evidence† | 9 | State the process for selecting sources of evidence (i.e., screening and eligibility) included in the scoping review. | 16-17 |
| Data charting process‡ | 10 | Describe the methods of charting data from the included sources of evidence (e.g., calibrated forms or forms that | 17-20 |

| | | | |
|---|---|---|---|
| | | have been tested by the team before their use, and whether data charting was done independently or in duplicate) and any processes for obtaining and confirming data from investigators. | |
| Data items | 11 | List and define all variables for which data were sought and any assumptions and simplifications made. | 20-22 |
| Critical appraisal of individual sources of evidence§ | 12 | If done, provide a rationale for conducting a critical appraisal of included sources of evidence; describe the methods used and how this information was used in any data synthesis (if appropriate). | N/A |
| Synthesis of results | 13 | Describe the methods of handling and summarizing the data that were charted. | 19 |
| **RESULTS** | | | |
| Selection of sources of evidence | 14 | Give numbers of sources of evidence screened, assessed for eligibility, and included in the review, with reasons for exclusions at each stage, ideally using a flow diagram. | 4 |
| Characteristics of sources of evidence | 15 | For each source of evidence, present characteristics for which data were charted and provide the citations. | 5-11 |
| Critical appraisal within sources of evidence | 16 | If done, present data on critical appraisal of included sources of evidence (see item 12). | N/A |
| Results of individual sources of evidence | 17 | For each included source of evidence, present the relevant data that were charted that relate to the review questions and objectives. | 5-11 |
| Synthesis of results | 18 | Summarize and/or present the charting results as they relate to the review questions and objectives. | 5-11 |
| **DISCUSSION** | | | |
| Summary of evidence | 19 | Summarize the main results (including an overview of concepts, themes, and types of evidence available), link to the review questions and objectives, and consider the relevance to key groups. | 12 |
| Limitations | 20 | Discuss the limitations of the scoping review process. | 14 |
| Conclusions | 21 | Provide a general interpretation of the results with respect to the review questions and objectives, as well as potential implications and/or next steps. | 14 |
| **FUNDING** | | | |
| Funding | 22 | Describe sources of funding for the included sources of evidence, as well as sources of funding for the scoping review. Describe the role of the funders of the scoping review. | 21 |

### Supplementary Material 2: Search Terms According to PCC Frame

| PCC Element | Category | Keywords |
|---|---|---|
| **Population** | **Dementia** | Alzheimer's disease, vascular dementia, Lewy body dementia, Parkinson's disease dementia, early-onset dementia, mixed dementia, frontotemporal dementia, unspecified dementia |
| | **Neurodegenerative Disease** | Mild cognitive impairment, Parkinson's disease, amyotrophic lateral sclerosis, motor neuron disease, Huntington's disease |
| **Concept** | (A) **Technology Types** | |
| | **Wearable** | Wearable sensors, wearable electronic devices, actigraphy, accelerometry, smartwatch, activity tracker, wristband, inertial measurement unit, IMU, gyroscope, wearable ECG, wearable EEG |
| | **Smart Home** | Smart home technology, ambient assisted living, passive infrared sensor, PIR, contact sensor, smart plug, environmental sensor, in-home monitoring, ambient sensor, smart home system |
| | **Telehealth** | Telehealth, telemedicine, telerehabilitation, videoconference, remote consultation, remote visit, video call, digital health platform, virtual clinic |
| | **Mobile App** | Mobile application, smartphone application, tablet application, mHealth app, digital app, mobile health, app-based intervention |
| | **Software** | Computer software, computer-based program, digital cognitive assessment, web-based platform, software tool, desktop application, computer-based intervention |
| | **Robotics** | Social assistive robot, SAR, robot, robotic device, socially assistive robotics, companion robot, care robot |
| | (B) **Technology Domains** | |
| | **Mobility** | Gait, postural balance, locomotion, fall detection, fall prevention, fall monitoring, fall risk, wander detection, wandering prevention, GPS tracking, geofencing, location tracking |
| | **Health Monitoring** | Vital signs, physiological monitoring, heart rate, blood pressure, sleep tracking, sleep quality, circadian rhythm, agitation, aggression, behavioural symptoms (BPSD), neuropsychiatric symptoms (NPS) |
| | **ADL Support** | Activities of daily living (ADL, IADL), self-care, bathing, dressing, eating, toileting, grooming, medication adherence, medication reminder, medication management, routine monitoring, activity recognition |
| | **Caregiver Support** | Caregiver burden, carer burden, caregiver support, caregiver stress, burnout, compassion fatigue, respite care, caregiver intervention, caregiver training, family member, spouse, informal caregiver |
| | **Communication** | Communication aid, communication device, social engagement, social isolation, loneliness, video call, videoconference, cognitive stimulation, cognitive engagement, reminder, memory aid, augmentative and alternative communication (AAC) |
| **Context** | **Setting** | Home, community, independent living, lab-based |

### Supplementary Material 3: Search Strategy

Database(s): Ovid MEDLINE(R) ALL 1946 to November 05, 2025

Search Strategy:
Searches Results

1 exp Dementia/ 232731
2 exp Alzheimer Disease/ 137104
3 exp Cognitive Dysfunction/ 51038
4 exp Parkinson Disease/ 92342
5 exp Motor Neuron Disease/ 37671
6 exp Huntington Disease/ 14612
7 dementia*.mp. 189116
8 alzheimer*.mp. 239878
9 (ADRD or "Alzheimer disease and related dementias").mp. 1834
10 "cognitive impair*".mp. 116632
11 ("mild cognitive impairment" or MCI).mp. 41082
12 (neurodegenerative adj3 disease*).mp. 101501
13 ("frontotemporal dementia" or FTD).mp. 12720
14 ("vascular dementia" or VaD).mp. 18496
15 ("lewy body" or "dementia with lewy bodies" or DLB or LBD or "parkinson* disease dementia" or PDD).mp. 18962
16 ("early onset dementia" or "young onset dementia" or EOD or YOD).mp. 2437
17 parkinson*.mp. 181187
18 ("motor neuron* disease*" or "motor neurone* disease*" or MND).mp. 12007
19 ("amyotrophic lateral sclerosis" or ALS).mp. 95666
20 ("huntington* disease*" or HD).mp. 69695
21 1 or 2 or 3 or 4 or 5 or 6 or 7 or 8 or 9 or 10 or 11 or 12 or 13 or 14 or 15 or 16 or 17 or 18 or 19 or 20 804700
22 exp Wearable Electronic Devices/ 24846
23 Actigraphy/ 5482
24 Accelerometry/ 9294
25 (wearable* adj3 (device* or sensor* or technolog*)).mp. 30599
26 (smartwatch* or "smart watch*").mp. 2209
27 ("fitness tracker*" or "activity tracker*").mp. 2720
28 (actigraph* or acceleromet*).mp. 38620
29 (wristband* or "smart band*" or "ring sensor*").mp. 1258
30 ("body-worn sensor*" or "bodyworn sensor*").mp. 260
31 ("inertial measurement unit*" or IMU).mp. 5350

32 22 or 23 or 24 or 25 or 26 or 27 or 28 or 29 or 30 or 31 83653
33 Remote Sensing Technology/ 5008
34 Telemedicine/ 46661
35 Monitoring, Ambulatory/ 8834
36 exp Environmental Monitoring/ 195115
37 (remote adj3 (monitor* or sens*)).mp. 29535
38 (telemonitor* or telehealth or telemedicine).mp. 70695
39 ("ambient sensor*" or "environmental sensor*").mp. 901
40 ("motion sensor*" or "PIR sensor*" or "passive infrared sensor*" or "occupancy sensor*").mp. 1721
41 ("smart home*" or "intelligent home*").mp. 1474
42 ("ambient assist* living" or AAL).mp. 1491
43 ("pressure sensor*" or "bed sensor*" or "chair sensor*").mp. 6171
44 ("camera-based" or "video-based" or "depth sensor*").mp. 7618
45 (radar or "radio frequency" or ultrasound or lidar).mp. 399473
46 33 or 34 or 35 or 36 or 37 or 38 or 39 or 40 or 41 or 42 or 43 or 44 or 45 708841
47 exp Self-Help Devices/ 14574
48 Self-Help Devices/ 6356
49 (assistive adj3 technolog*).mp. 4991
50 (assistive adj3 device*).mp. 4911
51 ("assistive technolog*" or "assistive device*").mp. 8530
52 ("GPS device*" or "tracking device*" or geofenc*).mp. 2397
53 ("wayfinding technolog*" or "navigation assist*" or "location monitor*").mp. 829
54 ("smart pill box*" or "medication dispenser*").mp. 71
55 ("home automation" or domotics or robotics or "internet of things" or IoT).mp. 57145
56 (robot* adj3 (social or companion or assistive or care)).mp. 2782
57 ("voice assistant*" or "virtual assistant*" or chatbot*).mp. 4146
58 ("digital companion*" or "AI assistant*" or "speech interface").mp. 191
59 ("mobile app*" or "smartphone app*" or "tablet computer*").mp. 30896
60 47 or 48 or 49 or 50 or 51 or 52 or 53 or 54 or 55 or 56 or 57 or 58 or 59 114501
61 32 or 46 or 60 880618
62 exp Gait/ 41967
63 Accidental Falls/ 30733
64 Postural Balance/ 31575
65 Locomotion/ 30064
66 (fall* adj3 (detect* or prevent* or monitor* or risk*)).mp. 27073
67 (gait adj3 (monitor* or analys* or pattern* or speed)).mp. 26026

68 (walk* adj3 (pattern* or monitor* or speed)).mp. 15871
69 (wander* adj3 (detect* or monitor* or prevent*)).mp. 106
70 ("GPS track*" or geofenc* or "location track*").mp. 1157
71 (mobility or locomotion or ambulation).mp. 303384
72 62 or 63 or 64 or 65 or 66 or 67 or 68 or 69 or 70 or 71 408535
73 "Activities of Daily Living"/ 77789
74 Self Care/ 38076
75 Medication Adherence/ 21546
76 ("activit* of daily living" or ADL or IADL).mp. 101424
77 ("self-care" or "self care").mp. 57892
78 (bathing or dressing or eating or toileting or grooming).mp. 223861
79 (medication adj3 (remind* or manag* or adherence or monitor*)).mp. 53120
80 ("routine monitor*" or "activity recognition").mp. 6474
81 73 or 74 or 75 or 76 or 77 or 78 or 79 or 80 431198
82 Vital Signs/ 2473
83 Heart Rate/ 183039
84 Blood Pressure/ 301549
85 exp Sleep/ 109925
86 ("vital sign*" or "physiologic* monitor*" or "physiological monitor*").mp. 27103
87 ("heart rate" or "blood pressure" or "pulse rate").mp. 692768
88 (sleep adj3 (track* or monitor* or quality or pattern*)).mp. 50936
89 ("circadian rhythm*" or "sleep-wake").mp. 131603
90 (agitation or aggression or "behavioral symptom*" or "behavioural symptom*" or BPSD).mp. 91481
91 ("neuropsychiatric symptom*" or NPS).mp. 80191
92 82 or 83 or 84 or 85 or 86 or 87 or 88 or 89 or 90 or 91 1096320
93 Caregiver Burden/ 1150
94 Caregivers/ 59631
95 Respite Care/ 1121
96 (caregiver* or carer*).mp. 146390
97 ("family member*" or spouse* or "adult child*").mp. 164573
98 ("caregiver burden" or "carer burden").mp. 7830
99 ("caregiver support" or "carer support" or "caregiver assist*" or "carer assist*").mp. 2339
100 ("caregiver stress" or "carer stress" or burnout or "compassion fatigue").mp. 36222
101 ("caregiver intervention*" or "carer intervention*" or "caregiver training" or "carer training").mp. 1081

102 ("respite care" or "respite service*" or "day care centre" or "day care center").mp. 3098
103 93 or 94 or 95 or 96 or 97 or 98 or 99 or 100 or 101 or 102 331983
104 Communication Devices for People with Disabilities/ 3217
105 Social Isolation/ 17820
106 Reminder Systems/ 4026
107 ("communication aid*" or "communication device*").mp. 4522
108 ("social engagement" or "social isolation" or loneliness).mp. 46208
109 ("video call*" or "video conferenc*" or videoconferenc* or "video chat*" or videophone*).mp. 9613
110 ("cognitive stimulation" or "cognitive engagement").mp. 2238
111 (reminder* or prompt* or cue*).mp. 382125
112 ("memory aid*" or "memory support*").mp. 747
113 ("assistive communication" or "augmentative communication" or AAC).mp. 7966
114 104 or 105 or 106 or 107 or 108 or 109 or 110 or 111 or 112 or 113 450401
115 72 or 81 or 92 or 103 or 114 2579530
116 Independent Living/ 14753
117 Home Care Services/ 38613
118 Community Health Services/ 34015
119 (home* or "home-based" or "in-home").mp. 838287
120 ("community dwelling" or "community living").mp. 41049
121 ("aging in place" or "ageing in place").mp. 1576
122 "independent living".mp. 18008
123 ("community setting*" or "non-institutional*").mp. 17362
124 116 or 117 or 118 or 119 or 120 or 121 or 122 or 123 921686
125 21 and 61 and 115 and 124 1613
126 limit 125 to english language 1580

### Supplementary Table S1: Study characteristics across primary diagnostic groups

Sample size = cognitive + motor + caregiver groups (controls excluded). Duration: median of all studies with parseable duration, including sub-day designs. Custom/prototype = Custom-Built / Lab Prototype. Validated usability = SUS, UEQ, UTAUT, or equivalent.

| Primary diagnosis | n | % of 325 | Sample size, median (IQR) | Duration, median d | Custom/prototype hardware | Validated usability tool |
|---|---|---|---|---|---|---|
| Parkinson's disease | 161 | 49.5 | 23 (12-45) | 14 | 108 (67.1%) | 8 (5.0%) |
| Alzheimer's disease | 62 | 19.1 | 20 (10-35) | 56 | 41 (66.1%) | 7 (11.3%) |
| Other dementia subtypes | 47 | 14.5 | 34 (15-92) | 84 | 32 (68.1%) | 2 (4.3%) |
| Mild cognitive impairment | 23 | 7.1 | 15 (10-30) | 87 | 20 (87.0%) | 2 (8.7%) |
| Motor neuron disease / ALS | 13 | 4 | 19 (8-25) | 60 | 9 (69.2%) | 0 (0.0%) |
| Huntington's disease | 9 | 2.8 | 15 (12-20) | 8 | 4 (44.4%) | 0 (0.0%) |
| Mixed neurodegenerative | 10 | 3.1 | 54 (24-65) | 42 | 4 (40.0%) | 0 (0.0%) |

### Supplementary Table S2: Reported technical difficulties by technology domain

Multi-select extraction field; a single study may contribute to more than one issue. 155 of 325 studies (47.7%) did not report any technical issue (144 explicit 'No'; 11 unrecorded).

| Technical issue category | Mobility | Health Mon. | ADL Support | Caregiver Sup. | Communication | Total |
|---|---|---|---|---|---|---|
| Connectivity/Sync | 20 | 24 | 6 | 10 | 6 | 66 |
| Software Bug | 24 | 16 | 9 | 13 | 3 | 65 |
| Hardware Breakage | 9 | 14 | 5 | 4 | 2 | 34 |
| User Burden/Error | 12 | 12 | 2 | 2 | 2 | 30 |
| Technical Failure | 6 | 12 | 4 | 3 | 1 | 26 |
| Battery | 4 | 5 | 0 | 3 | 1 | 13 |
| Measurement/Method Limitation | 4 | 4 | 1 | 0 | 0 | 9 |
| Sensor Issues | 4 | 4 | 0 | 0 | 1 | 9 |
| Data Loss | 3 | 2 | 0 | 1 | 0 | 6 |

### Supplementary Table S3: Studies with N≥50 and evaluation duration >12 months

Nineteen studies (5.8% of 325) met both thresholds. Two Fleisher 2022 entries are distinct papers from the IN-HOME-PD programme (patient cohort vs. caregiver cohort).

| Covidence ID | First author | Year | Domain | n | Duration |
|---|---|---|---|---|---|
| 374 | Puaschitz et al. | 2021 | ADL Support | 552 | 24 Months |
| 376 | Robert Howard, Rebecca Gathercole, Rosie Bradley, et al. | 2021 | ADL Support | 495 | 104 Weeks |
| 379 | Rebecca Gathercole, Rosie Bradley, Emma Harper, Lucy Davies, Lynn Pank, Natalie Lam, Anna Davies, Emma Talbot, Emma Hooper, Rachel Winson, Bethany Scutt, Victoria Ordonez Montano, Samantha Nunn, Grace Lavelle, Matthew Lariviere, Shashivadan Hirani, Stefano Brini, Andrew Bateman, Peter Bentham, Alistair Burns, Barbara Dunk, Kirsty Forsyth, Chris Fox, Catherine Henderson, Martin Knapp, Iracema Leroi, Stanton Newman, John OBrien, Fiona Poland, John Woolham, Richard Gray and Robert Howard | 2021 | ADL Support | 990 | 104 Weeks |
| 601 | Michael C B David, Magdalena Kolanko, Martina Del Giovane, Helen Lai, Jessica True, Emily Beal, Lucia M Li, Ramin Nilforooshan, Payam Barnaghi, Paresh A Malhotra, Helen Rostill, David Wingfield, Danielle Wilson, Sarah Daniels, David J Sharp, Gregory Scott | 2023 | Health Monitoring | 164 | 15.8 To 1077.1 Days |
| 604 | Bafaloukou, M. et al. | 2025 | Health Monitoring | 112 | 3 Years |
| 1198 | Joseph E. Gaugler, Rachel Zmora, Lauren L. Mitchell, Jessica Finlay, Christina E. Rosebush, Manka Nkimbeng, Zachary G. Baker, Elizabeth A. Albers and Colleen M. Peterson | 2021 | Caregiver Support | 358 | 18 Months |
| 1511 | D M Feeney Mahoney et al. | 2001 | Caregiver Support | 142 | 18 Months |
| 2377 | Nan Fletcher-Lloyd, Alina-Irina Serban, Magdalena Kolanko, David Wingfield, Danielle Wilson, Ramin Nilforooshan, Payam Barnaghi, and Eyal Soreq | 2023 | Health Monitoring | 73 | 499 Days |
| 3037 | Skeldon et al. | 2025 | Health Monitoring | 70 | 2 Years |
| 3146 | Bahareh Chimehi, Julien Larivière-Chartier, Bruce Wallace, Zachary Beattie, Laura Ault, Lyndsey Miller, Joel Steele, Neil Thomas | 2023 | Caregiver Support | 94 | 700 Days |
| 4764 | Riikonen, M. et al. | 2013 | ADL Support | 50 | 3 Years |
| 13603 | Alireza Parsay, Mert Torun, Philip R. Delio, and Yasamin Mostofi | 2025 | Mobility | 68 | 1 Year |

| | | | | | |
|---|---|---|---|---|---|
| 18417 | Bart R. Maas et al. | 2024 | Health Monitoring | 200 | 1 Year |
| 18575 | Silva de Lima et al. | 2019 | Mobility | 2063 | 2.5 Years |
| 18902 | Jori E. Fleisher, Serena P. Hess, Ellen C. Klostermann, Jeanette Lee, Erica Myrick, Daniela Mitchem, Claire Niemet, Katheryn Woo, Brianna J. Sennott, Maya Sanghvi, Natalie Witek, James C. Beck, Jayne R. Wilkinson, Bichun Ouyang, Deborah A. Hall, and Joshua Chodosh | 2022 | Health Monitoring | 130 | 1 Year |
| 19073 | Memedi M et al. | 2015 | Health Monitoring | 65 | 3 Years |
| 19214 | Soreq et al. | 2025 | Health Monitoring | 83 | 6 Years |
| 19805 | Jori E. Fleisher, Madhuvanthi Suresh, Ellen C. Klostermann, Jeanette Lee, Serena P. Hess, Erica Myrick, Daniela Mitchem, Katheryn Woo, Brianna J. Sennott, Natalie Witek, Sarah Mitchell Chen, James C. Beck, Bichun Ouyang, Jayne R. Wilkinson, Deborah A. Hall, and Joshua Chodosh | 2022 | Caregiver Support | 130 | 1 Year |
| 29331 | Burq et al. | 2022 | Mobility | 388 | 70 Weeks |

### Supplementary Table S4: Evaluation duration by primary diagnostic group

| **Primary Diagnosis** | **n (total)** | **n (day-or-longer)** | **n (sub-day/sessionless)** | **Duration Days, Median (IQR)** | **Max Duration (days)** |
|---|---|---|---|---|---|
| Parkinson's disease | 161 | 121 | 40 | 14 (5-56) | 1095 |
| Alzheimer's disease | 62 | 49 | 13 | 56 (16-150) | 2190 |
| Other dementia subtypes | 47 | 44 | 3 | 84 (30-180) | 728 |
| Mild cognitive impairment | 23 | 14 | 9 | 87 (36-148) | 360 |
| Motor neuron disease / ALS | 13 | 11 | 2 | 60 (56-168) | 360 |
| Huntington's disease | 9 | 9 | 0 | 8 (7-60) | 360 |
| | 10 | 9 | 1 | 42 (3-180) | 730 |

### Supplementary Table S5: Sub-Day and Single-Session studies subset

| | |
|---|---|
| **Overall Characteristics** | |
| Total studies in subset | **68** |
| Sessionless (no parseable duration) | **1** |
| Sub-day (parseable, <1 day) | **67** |
| % of 325 included studies | **20.9** |
| Median sample size | **20** |
| Sample size IQR | **10-30** |
| % custom-built / prototype | **89.7** |
| % using validated usability instrument | **2.9** |
| **By primary domain** | |
| Mobility | **35** |
| Health Monitoring | **12** |
| ADL Support | **14** |
| Caregiver Support | **6** |
| Communication | **1** |
| **By study design** | |
| Active | **14** |
| Passive | **35** |
| Mixed/Other | **19** |
| **By primary diagnosis** | |
| Parkinson's disease | **40** |
| Alzheimer's disease | **13** |
| Other dementia subtypes | **3** |
| Mild cognitive impairment | **9** |
| Motor neuron disease / ALS | **2** |
| Mixed neurodegenerative | **1** |
| Huntington’s disease | **0** |

### Supplementary Figure 1

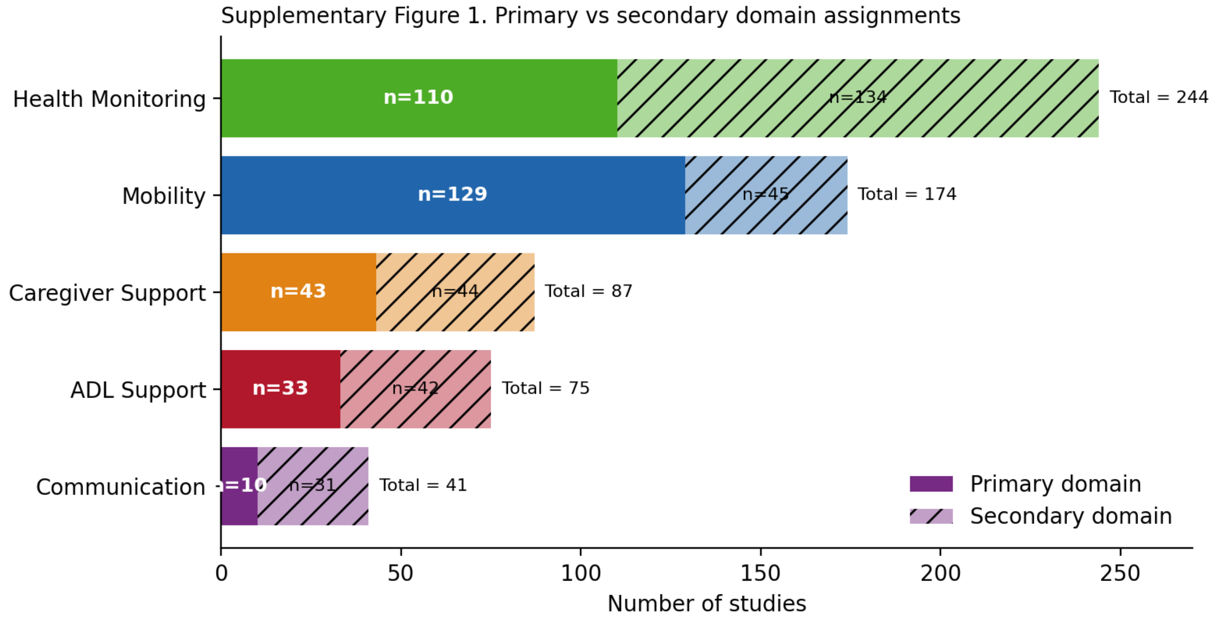


### Supplementary Figure 2

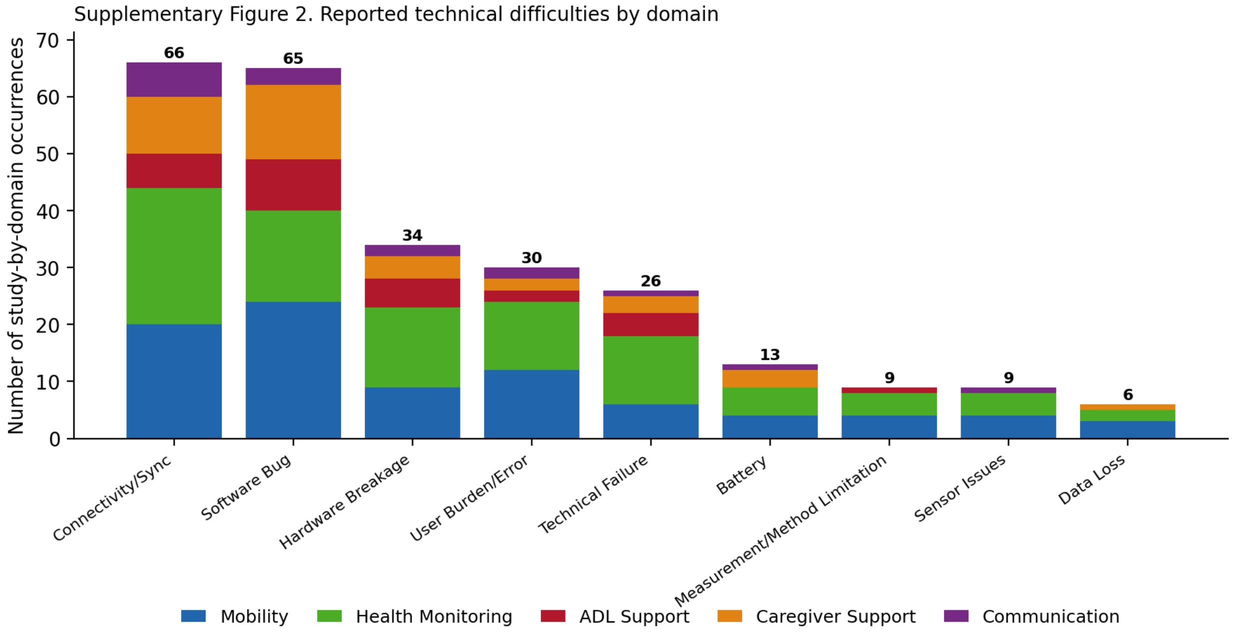
